# Simulating desegregation through affordable housing development: an environmental health impact assessment of Connecticut zoning law

**DOI:** 10.1101/2024.02.13.24302645

**Authors:** Saira Prasanth, Nire Oloyede, Xuezhixing Zhang, Kai Chen, Daniel Carrión

## Abstract

Residential segregation shapes access to health-promoting resources and drives health inequities in the United States. Connecticut’s Section 8-30g incentivizes municipalities to develop a housing stock that is at least 10% affordable housing. We used this implicit target to project the impact of increasing affordable housing across all 169 Connecticut municipalities on all-cause mortality among low-income residents. We modeled six ambient environmental exposures: fine particulate matter (PM_2.5_), ozone (O_3_), nitrogen dioxide (NO_2_), summertime daily maximum heat index, greenness, and road traffic noise. We allocated new affordable housing to reach the 10% target in each town and simulated random movement of low-income households into new units using an inverse distance weighting penalty. We then quantified exposure changes and used established exposure-response functions to estimate deaths averted stratified by four ethnoracial groups: Asian, Hispanic or Latino, non-Hispanic Black, and non-Hispanic White. We quantified racialized segregation by computing a multi-group index of dissimilarity at baseline and post-simulation. Across 1,000 simulations, in one year (2019) we found on average 169 (95% CI: 84, 255) deaths averted from changes in greenness, 71 (95% CI: 49, 94) deaths averted from NO_2_, 9 (95% CI: 4, 14) deaths averted from noise, and marginal impacts from other exposures, with the highest rates of deaths averted observed among non-Hispanic Black and non-Hispanic White residents. Multi-group index of dissimilarity declined on average in all eight Connecticut counties post-simulation. Sensitivity analyses simulating a different population movement strategy and modeling a different year (2018) yielded consistent results. Strengthening desegregation policy may reduce deaths from environmental exposures among low-income residents. Further research should explore non-mortality impacts and additional mechanisms by which desegregation may advance health equity.

## 1. Introduction

Residential segregation has a profound impact on health and well-being. Segregation is associated with inequities in preterm birth, infectious disease, cancer, asthma, mental health, and other health outcomes (Anderson et al., 2021; Krieger et al., 2020; Nardone et al., 2020). In the United States, residential segregation is rooted in a history of exclusionary zoning and lending practices. This is most emblematic via “redlining,” a practice in 1930-1934 through which the Home Owners’ Loan Corporation designated predominantly Black or low-wealth neighborhoods as “hazardous” for mortgage investment while grading predominantly White or affluent neighborhoods as “best”, reinforcing racialized economic segregation (Krieger et al., 2020; Nardone et al., 2020; Nardone et al., 2021). Despite the 1968 Fair Housing Act, segregation is far from declining - a recent report found that 54% of U.S. metropolitan regions were more highly segregated in 2020 than in 1990 (Menendian et al., 2021; Steil et al., 2021).

Residential segregation shapes access to housing, wealth, education, nutrition, and other aspects of the contextual environment integral to long-term health (Appel & Nickerson, 2016; Eaton, 2020; Havewala, 2021; Menendian et al., 2021). Environmental racism meanwhile describes the placement of adverse exposures (e.g., pollution, extreme heat, noise) and deprivation of environmental goods (e.g., greenspace) among residents who are Black, Indigenous, Hispanic or Latino (herein referred to as Latino), and other People of Color (Carrión et al., 2022; Seamster & Purifoy, 2020). Consequently, residential segregation represents collocation of adverse social and environmental exposures in neighborhoods with predominantly low-wealth, Black, or Latino residents, as well as concentration of health-promoting resources in neighborhoods with predominantly White or affluent residents (Carrión et al., 2022; Seamster & Purifoy, 2020).

Integrative policies are the most likely means to address environmental injustice. Presently, single-family zoning and other exclusionary tools that privilege low-density housing stall development of affordable multifamily units in areas of accumulated wealth and social capital, where “not in my backyard” opposition is often most potent (Girouard, 2023; Lens, 2022; Steil & Lens, 2023; Whittemore, 2020). These ordinances frequently push the burden of development onto neighborhoods impacted by disinvestment and made socially vulnerable to gentrification (Whittemore, 2020). Dismantling exclusionary zoning and equitably expanding housing supply meanwhile remain critical to reduce housing cost burden and housing instability, which disproportionately impact low-income and minoritized residents (Flint, 2022; Harvard Law Review, 2022; Lens, 2022; Swope & Hernández, 2019). This densification, alongside other zoning interventions such as transit-oriented development, also has promising environmental co-benefits (Harvard Law Review, 2022). Yet, several recent attempts to legislate sweeping zoning reforms - including stronger reforms proposed alongside Connecticut’s recently passed Public Act 21-29 - have been abrogated or diluted by opponents of state zoning preemption (Flint, 2022; Harvard Law Review, 2022).

Section 8-30g of the Connecticut General Statutes provides a case study of one existing state-level place-based policy that aims to equitably increase affordable housing supply (CT Gen Stat, 2022; Desegregate Connecticut, n.d. -a; Steil & Lens, 2023). Connecticut is highly segregated - two-thirds of Black and Latino residents lived in only 15 out of 169 municipalities in 2010 (Eaton, 2020). Section 8-30g, passed in 1989, established the Affordable Housing Land Use Appeals Procedure, through which developers can appeal local zoning decisions that deny or significantly restrict affordable housing proposals (CT Gen Stat, 2022; Desegregate Connecticut, n.d. -a). Contrary to the status quo, Section 8-30g shifts the burden of proof in these appeals off developers and onto municipalities, which must prove that the decision is grounded in public health or other valid considerations *and* that these issues outweigh the need for affordable housing (CT Gen Stat, 2022; Desegregate Connecticut, n.d. -a). Municipalities have won only about one-third of Section 8-30g appeals (Desegregate Connecticut, n.d. -a).

Additionally, Section 8-30g exempts municipalities if at least 10% of their housing stock is affordable, or if they qualify for a moratorium by demonstrating adequate progress toward this target (CT Gen Stat, 2022; Desegregate Connecticut, n.d. -a). Despite this incentive, only 29 out of 169 Connecticut municipalities met the 10% benchmark in 2022 (Zaldonis & Connecticut Department of Housing, 2023). Therefore, an important policy question remains as to whether full realization of the implicit 10% target can advance desegregation and environmental health equity.

The aim of this study was to assess potential mortality reductions among low-income Connecticut residents from strengthening desegregation policy to enforce the 10% benchmark across all townships. We quantified mortality impacts in terms of all-cause deaths averted from six ambient environmental exposures, stratified by ethnoracial group. We hypothesized that desegregation policy implementation would avert mortality among low-income households across all ethnoracial groups from changes in each of the following ambient exposures: fine particulate matter (PM_2.5_), ozone (O_3_), nitrogen dioxide (NO_2_), summertime daily maximum heat index, greenness, and road traffic noise. We also hypothesized that the rate of all-cause deaths averted would be higher among non-Hispanic Black and Latino low-income households compared to non-Hispanic White and Asian low-income households.

## 2. Methods

We simulated random movement of low-income households into hypothetical new affordable housing and used published exposure–response functions to estimate mortality impacts over one year associated with each environmental exposure, stratified by ethnoracial group.

### 2.1. Environmental data

Our analysis focused solely on exposures in the ambient rather than indoor environment because we do not have access to indoor environmental exposure models. We retrieved daily average PM_2.5_ levels and daily 8-hour maximum O_3_ levels by census tract centroid in 2019 from the U.S. Environmental Protection Agency’s Fused Air Quality Surface Using Downscaling datasets (U.S. Environmental Protection Agency, 2023). Daily values were averaged across the year and across tracts using areal-weighted interpolation by township (**Figure A.1.a,b**). NO_2_ annual averages were retrieved from NASA’s Nitrogen Dioxide Surface-Level Annual Average Concentrations V1 dataset in 0.0083 degree gridded estimates (**Figure A.1.c**) (Anenberg, 2023). Gridded 1 x 1-km daily maximum air temperature and vapor pressure from NASA’s Daymet datasets were used to calculate daily maximum heat index (**Figure A.1.d**) (Anderson et al., 2013; Thornton et al., 2022; Thornton et al., 2000; Thornton & Running, 1999; Thornton et al., 1997; Thornton et al., 2021). Additionally, we retrieved 1 x 1-km gridded estimates of monthly average normalized difference vegetation index (NDVI) from NASA’s Terra MODIS Vegetation Indices (MOD13A3.061) (**Figure A.1.e**) (Didan, 2021). Finally, 30 x 30-m gridded road traffic noise estimates in 2018 and 2020 were retrieved from the Bureau of Transportation Statistics National Transportation Noise Map, and linear imputation was used to estimate 2019 values (**Figure A.1.f**) (U.S. Department of Transportation, 2022). Population-weighted annual or daily averages were derived for all gridded exposure estimates (NO_2_, heat, NDVI, noise) by township using the Columbia University Center for International Earth Science Information Network’s 2020 Gridded Population of the World (Center for International Earth Science Information Network, 2018).

### 2.2. Housing, demographic, and health data

We retrieved counts of affordable and total housing units by township from the Affordable Housing Land Use Appeals List through Connecticut Open Data (**Figure A.2.a,b**) (Zaldonis & Connecticut Department of Housing, 2023). Households by income bracket, town, and ethnoracial group (Asian, Latino, non-Hispanic Black, and non-Hispanic White) and average household size by town were retrieved from 2020 Decennial Census and 2019 American Community Survey five-year estimates (U.S. Census Bureau, 2020, 2022a, 2022b, 2022c, 2022d, 2022e). Low-income status was assigned using the Department of Housing and Urban Development fiscal year 2019 three-person household income limits, based on statewide average household size (U.S. Census Bureau, 2022a; U.S. Department of Housing and Urban Development, 2023). Finally, we retrieved statewide crude all-cause mortality rates per 100,000 people by ethnoracial group, year, and month from the Centers for Disease Control and Prevention Wide-ranging Online Data for Epidemiologic Research (CDC WONDER) database (National Center for Health Statistics, 2022).

### 2.3. Simulating housing development and population movement

We first allocated development of new affordable housing units to each township so that a resulting 10% of total units were affordable units. Affordable housing as defined by the Department of Housing includes government-assisted housing, housing with tenants receiving rental assistance, housing financed by single-family Connecticut Housing Finance Authority and/or U.S. Department of Agriculture mortgages, and housing with deed restrictions pricing them as affordable for households earning 80% or less of the area median income (AMI) (Zaldonis & Connecticut Department of Housing, 2023).

We simulated random movement or non-movement of low-income households across towns into newly developed affordable housing. Low-income households were defined as households earning an annual income at or below 80% AMI in fiscal year 2019 in the HUD Metropolitan Fair Market Rent/Income Limits Area (HMFA) of residence (U.S. Department of Housing and Urban Development, 2023). We restricted households from moving to HMFAs where they would no longer fall at or below 80% AMI post-move.

We drew a random number of households for each possible pre/post-move town pair from a binomial distribution, where the mean count was weighted by availability of and proximity to new housing. Availability was measured by the proportion of new units built in the post-move town out of all units built. We then used an inverse distance weighting penalty to model the assumption that households were likely to preferentially move closer to their original home (Gillespie, 2017). Detailed weighting methods are described in **Text A.1**.

### 2.4. Assessing mortality impacts

For each household, we quantified changes in exposure to summertime daily maximum heat index and annual average PM_2.5_, O_3_, NO_2_, NDVI, and road noise at the township level. We used the following exposure–response functions to project changes in all-cause mortality associated with changes in each exposure:

- Hazard ratio of 1.12 (95% confidence interval: 1.08, 1.15) per 10 μg/m^3^ increase in long-term PM_2.5_ (Pope et al., 2019)
- Hazard ratio of 1.02 (1.01, 1.04) per 10 ppb increase in long-term O_3_ (Turner et al., 2016)
- Hazard ratio of 1.06 (1.04, 1.08) per 10 ppb increase in long-term NO_2_ (Huang et al., 2021)
- Rate ratio of 1.014 (1.004, 1.024) per 5° F increase in daily maximum heat index, between 75° F and 105° F (Wellenius et al., 2017)
- Hazard ratio of 0.96 (0.94, 0.97) per 0.1 increase in NDVI within 500 m (Rojas-Rueda et al., 2019)
- Risk ratio of 1.05 (1.02, 1.07) per 10 dB increase in road traffic noise (Hao et al., 2022)

In each simulation repeat, we randomly drew an effect estimate from a Normal distribution centered at the rate ratio for daily maximum heat index or the natural log of the hazard or risk ratio for PM_2.5_, O_3_, NO_2_, NDVI, and noise, with a standard deviation equal to the published margin of error divided by 1.96.

We calculated the attributable fraction for each annual exposure and for each pre-move (ⅈ) and post-move (*j*) town pair as follows, based on the pre/post change in exposure (*exposure*_*j*_ − *exposure*_*i*_), divided by the exposure change (*x*) corresponding to the function (10 units for PM_2.5_, O_3_, NO_2_, and noise; 0.1 unit for NDVI), and the randomly drawn coefficient for a given run (*k*) of the simulation (log(*HR*)_*k*_):

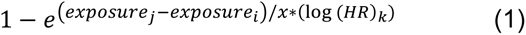

For daily maximum heat index, we calculated the attributable fraction for each day as follows, using the randomly drawn rate ratio for a given run of the simulation (*RR*_*k*_) corresponding to a 5° F increase:

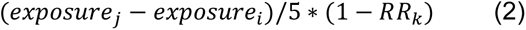

We then multiplied the attributable fraction by the number of households experiencing a given move, average household size in the pre-move town, and crude all-cause annual or daily mortality rate by ethnoracial group to estimate deaths averted in each potential move (Zocchetti, 2022). These were summed across all moves and, for daily deaths averted from heat, across all days in May 1–September 30, to estimate total deaths averted in one year from each exposure. We repeated this simulation 1,000 times to derive average estimates and 95% confidence intervals for deaths averted and rates of deaths averted per 100,000 low-income residents.

### 2.5. Segregation indices

We computed multi-group Duncan’s dissimilarity index at baseline and in the counterfactual population distribution to assess changes in the magnitude of segregation (Sakoda, 1981; Tivadar, 2019). We estimated this index at the county level, since dissimilarity index relies on summing population proportions in smaller administrative units (townships) across larger units (counties) (Sakoda, 1981; Tivadar, 2019).

### 2.6. Sensitivity analyses

First, we tested the sensitivity of our results to the inverse distance weighting penalty. Given that all eight Connecticut counties are considered one commuting zone, it is plausible that intra-state distances would not restrict movement to the extent assumed in the primary analysis (U.S. Department of Agriculture, 2012; Fowler & Jensen, 2020). We thus reproduced the analysis treating all movement distances within Connecticut as equally probable by removing our inverse distance weighting. Second, we tested the sensitivity of our results to the year of analysis by running the simulation using exposure, population, housing, income limit, and mortality data from 2018 instead of 2019.

### 2.7. Computation

All analyses were conducted using R version 4.2.1. R packages used for data retrieval, cleaning, and analysis are specified in **Text A.2** (Anderson et al., 2013; Hufkens et al., 2018; Richardson, 2022; Tivadar, 2019; Walker et al., 2024; Walker & Rudis, 2024).

## 3. Results

In 2019, we estimated there were 1,345,504 households in Connecticut with Asian, Latino, non-Hispanic Black, or non-Hispanic White householders. Of these households, 40.0% (n = 537,763) were designated as low-income. Across 1,000 simulations, on average, 39.6% of low-income households stayed in their town of residence, while 60.4% (approximately 324,823 households) moved to a different town. Total low-income households at baseline and post-simulation by township are shown in **Figure 1**, while low-income households by ethnoracial group at baseline and post-simulation are shown in **Figures A.3–A.6**.

**Figure 1.**
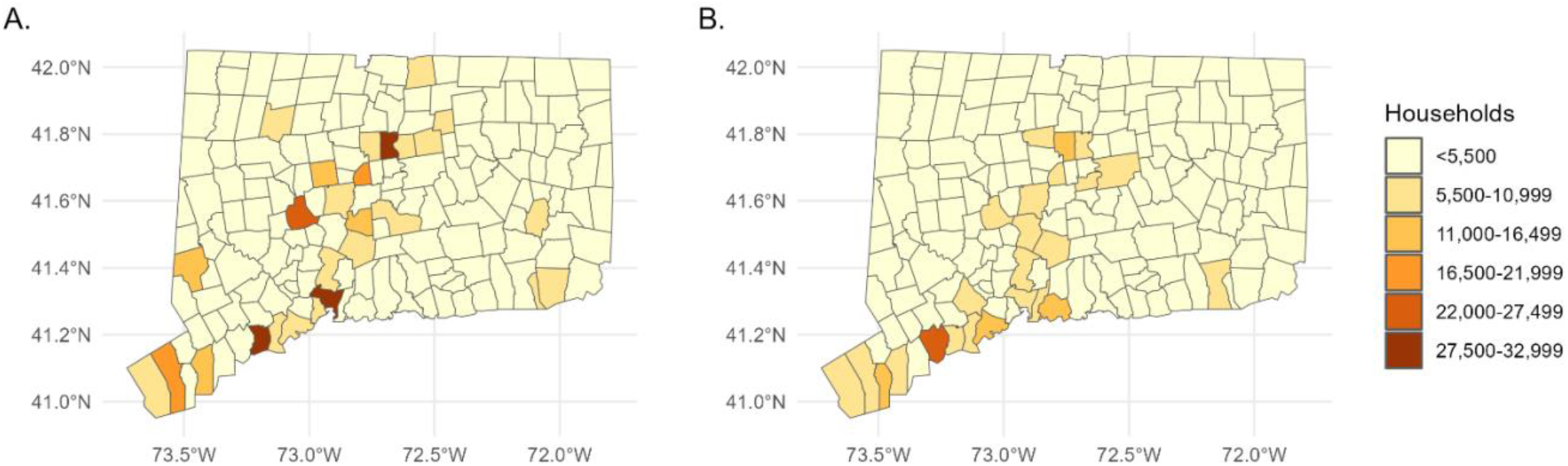
Map of low-income households by township A) at baseline in 2019 and B) post-move in 2019, averaged over 1,000 simulations.^a^ ^a^ Please use color for Figure 1 and appendix figures in print.

### 3.1. Index of dissimilarity

The simulation dispersed an average of 324,823 low-income households, most notably from towns with the highest concentrations of low-income households at baseline (e.g., Hartford, New Haven, Bridgeport) (**Figure 1**). Multi-group index of dissimilarity declined in all eight Connecticut counties after simulating desegregation, with the largest reduction in Middlesex County (**Table A.1, Figure A.7**).

### 3.2. Environmental exposures

Average town-level annual exposure to PM_2.5_ and O_3_ increased slightly post-simulation (**Table A.2**). We observed reductions in annual average NO_2_ exposure, summertime heat index, and road noise as well as an increase in annual average NDVI exposure (**Table A.2**).

### 3.3. Averted all-cause mortality

Deaths averted within each ethnoracial group were primarily driven by changes in exposure to greenness, followed by NO_2_ and noise (**Table 1**). The highest numbers of deaths averted from NDVI, NO_2_, and noise were among non-Hispanic White low-income residents. Asian low-income residents experienced marginal reductions in deaths from NDVI and NO_2_ and no change in deaths from noise, heat, O_3_, or PM_2.5_.

**Table 1.** Average deaths averted (95% confidence intervals) in Connecticut in 2019 across 1,000 simulated moves and corresponding exposure contrasts.

| **Ethnoracial group** | **Heat** | **NDVI** | **NO<sub>2</sub>** | **Noise** | **O<sub>3</sub>** | **PM<sub>2.5</sub>** |
| --- | --- | --- | --- | --- | --- | --- |
| Asian | 0 (0, 0) | 1 (1, 2) | 1 (0, 1) | 0 (0, 0) | 0 (0, 0) | 0 (0, 0) |
| Hispanic/Latino | 0 (0, 0) | 25 (12, 38) | 10 (7, 14) | 1 (0, 2) | 0 (0, 0) | 0 (0, 0) |
| Non-Hispanic Black | 0 (0, 1) | 40 (20, 60) | 17 (12, 23) | 1 (0, 1) | 0 (0, 0) | 1 (0, 1) |
| Non-Hispanic White | 1 (0, 1) | 103 (51, 155) | 43 (30, 57) | 7 (3, 11) | -1 (-1, 0) | -3 (-4, -2) |
| Total | 1 (0, 2) | 169 (84, 255) | 71 (49, 94) | 9 (4, 14) | -1 (-2, -1) | -2 (-3, -1) |

NDVI, normalized difference vegetation index.

Changes in mortality from PM_2.5_, O_3_, and heat were marginal; we observed on average two excess deaths from PM_2.5_, one excess death from O_3_, and one averted death from heat in total, but no change among Asian or Latino low-income households from these exposures and no change among non-Hispanic Black low-income households from O_3_ or heat (**Table 1**).

Average rates of deaths averted from NDVI and NO_2_ per 100,000 low-income residents were highest among non-Hispanic Black residents, while the rate of deaths averted from noise was highest among non-Hispanic White residents (**Table 2**). Rates of averted or excess deaths were lowest in magnitude among Asian residents across all exposures (**Table 2**).

**Table 2.**
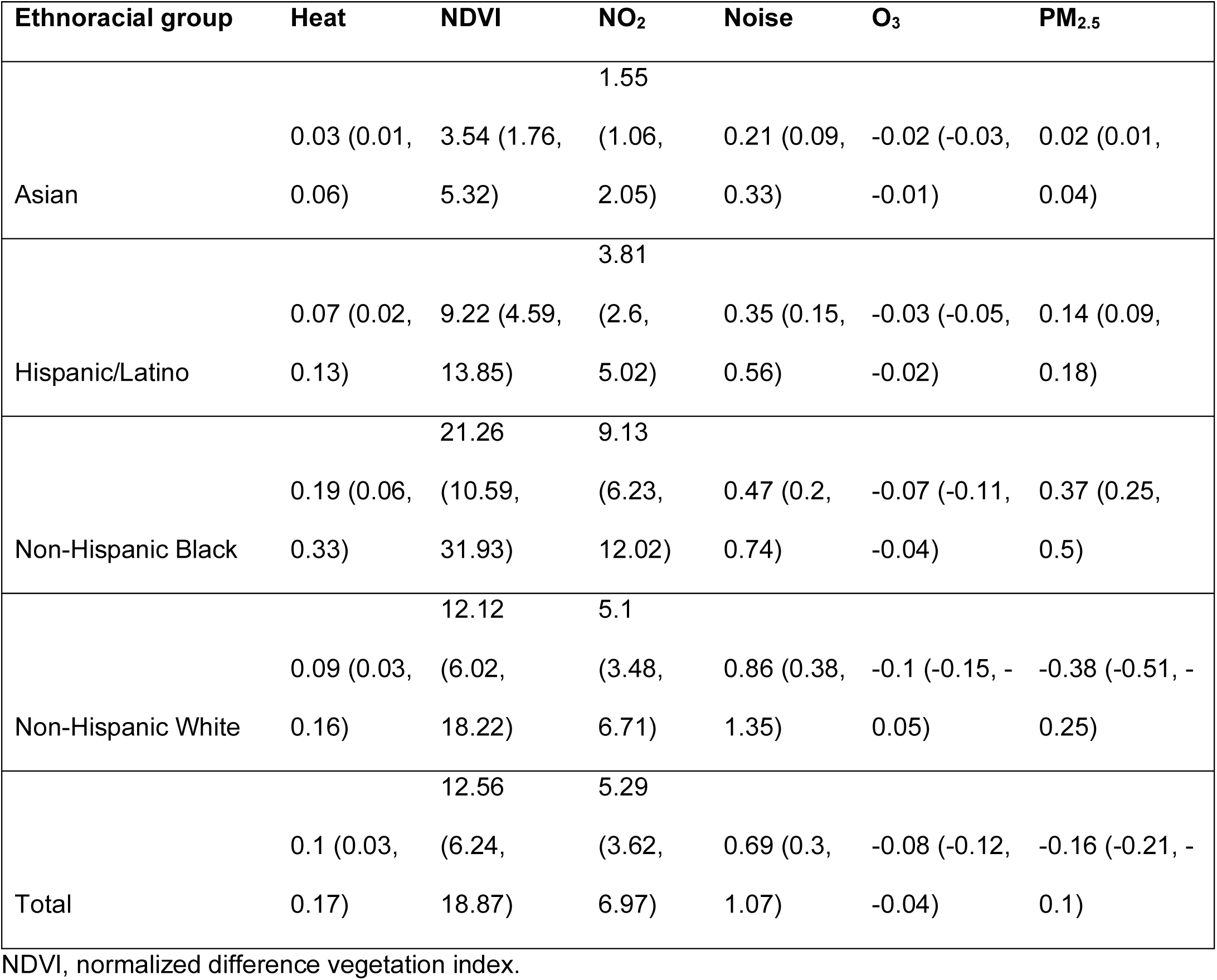
Rates of average deaths averted (95% confidence intervals) per 100,000 low-income residents in Connecticut in 2019 across 1,000 simulated moves and corresponding exposure contrasts.

### 3.4. Sensitivity analyses

Our first sensitivity analysis removed the inverse distance weighting penalty (**Table A.3**) and found that total deaths averted from NDVI, NO_2_, noise, and heat were slightly larger compared to deaths averted when movement was restricted according to distance (**Table 1**). Marginal averted deaths rather than excess deaths were associated with PM_2.5_, while excess deaths from O_3_ increased slightly (**Table A.3**).

Average deaths averted from NDVI in 2018 (**Table A.4**) were higher than 2019 estimates (**Table 1**). Deaths averted from noise were slightly higher and excess deaths from PM_2.5_ were slightly lower in 2018 simulations than in 2019, while impacts from other exposures remained constant (**Table A.4**).

## 4. Discussion

Our results suggest that full implementation of Section 8-30g’s 10% affordable housing target would reduce segregation in Connecticut and reduce mortality among low-income Connecticut residents. These results also suggest that the mortality benefits from desegregation policies would result primarily from increased exposure to greenness and reduced exposure to ambient NO_2_ and road noise. These benefits were most pronounced among non-Hispanic Black residents for NDVI and NO_2_, in line with our hypothesis that ethnoracial groups disproportionately impacted by environmental racism would experience the highest rates of averted deaths. Contrary to our hypothesis, the highest rate of averted deaths from noise, as well as the second highest rates of averted deaths from NDVI and NO_2_, were observed among non-Hispanic White residents, rather than non-Hispanic Black or Latino residents. We observed marginal changes in mortality from PM_2.5_, O_3_, and heat, suggesting that these exposure contrasts were not large enough to impact all-cause mortality. Our sensitivity analyses meanwhile suggest that mortality impacts remained consistent regardless of the year and thus may persist.

Strengthening Section 8-30g to more heavily incentivize or require the 10% benchmark presents an opportunity to advance environmental health justice by redistributing access to health-promoting environments while addressing the urgent need for housing. There are only an estimated 37 affordable and available housing units for every 100 extremely low-income renter households in Connecticut (National Low Income Housing Coalition, 2023). Meanwhile, recent polling reveals that 70 percent or more of Americans support policies to allow increased development of apartments (Horowitz & Kansal, 2023). Stronger state laws like Massachusetts’s Chapter 40B and California’s Senate Bill 9 may serve as models for strong zoning reform in Connecticut, which to date has been stymied in the legislature (Reid et al., 2017; California Senate Democratic Caucus, 2021; Flint, 2022; Harvard Law Review, 2022). For example, Connecticut’s recent Public Act 21-29 - which allows accessory dwelling units as-of-right in single-family residential zones, reduces parking space minimums, and mandates municipalities to submit plans to further affordable housing development - was arguably the weakest among three Connecticut zoning reform bills proposed in 2021 (Public Act 21-29, 2022; Desegregate Connecticut, n.d. -b; Harvard Law Review, 2022). Meanwhile, our finding of significant mortality impacts associated with exposure to NO_2_, a major traffic-related pollutant, and road noise highlights the ongoing racialized harms of highway construction in the U.S.

(Archer, 2020; Collins et al., 2020; Willis et al., 2023). Using the Federal Aid Highway Act of 1956, the U.S. government systematically demolished community institutions and displaced residents in predominantly Black neighborhoods, reinforcing residential segregation and exposure inequities (Archer, 2020; Collins et al., 2020; Willis et al., 2023). These policies appear to have reverberating impacts today.

By examining the implicit 10% affordable housing goal established by Section 8-30g, this analysis presents a case study of the potential health impacts of one place-based desegregation policy as well as a template for further state, regional, and national analyses. The need for legislative action toward health equity and the popularity of policies to increase apartment production underscore the importance of further research to evaluate actionable policy interventions (Horowitz & Kansal, 2023). Persistent reproduction of racialized health inequities, as well as the harm inflicted to date by extractive research practices and misrepresentation of race as a biological rather than social construct, moreover compel us to adopt action-oriented frameworks, center community expertise, and allocate research efforts to drive and strengthen solutions (Carrión et al., 2022; Lett, Adekunle, et al., 2022; Lett, Asabor, et al., 2022).

To our knowledge, this is the first environmental health analysis to simulate the potential role of zoning and housing policies toward mitigating segregation and exposure disparities. By using both availability of and proximity to new affordable housing to estimate movement probabilities, we simulated two major drivers of movement decisions (Gillespie, 2017). Our sensitivity analyses meanwhile suggest that our results are robust to assumptions about population movement and that our main results may be slightly conservative. A key strength of this health impact assessment is its consideration of six environmental exposures, each independently associated with mortality (Hao et al., 2022; Huang et al., 2021; Pope et al., 2019; Rojas-Rueda et al., 2019; Turner et al., 2016; Wellenius et al., 2017). Another strength is our analysis of mortality impacts stratified by ethnoracial group, highlighting the potential for this and similar policy interventions to advance environmental health equity.

However, there are limitations to this study. Our analysis reflected township-level zoning decisions, but this may have masked within-township exposure disparities. Additionally, our ethnoracial categorizations did not include Indigenous households due to low sample sizes and our inadequate power to detect an effect in this group. This remains pervasive and problematic among analyses of health effects across ethnoracial groups (Lett, Asabor, et al., 2022). We also assessed impacts using only one year and thus did not account for longer-term effects and annual variation in between-township exposure contrasts, although our results were robust to the year of choice in sensitivity analyses. Moreover, while we focused on environmental exposures and mortality, these policies would likely impact other health-relevant social exposures and non-mortality endpoints as well (Anderson et al., 2021; Appel & Nickerson, 2016; Eaton, 2020; Havewala, 2021; Krieger et al., 2020; Menendian et al., 2021; Nardone et al., 2020; Steil et al., 2021). Meanwhile, the exposure-response functions used in this analysis were not derived from studies specific to low-income Connecticut residents, potentially limiting their generalizability to our study population (Hao et al., 2022; Huang et al., 2021; Pope et al., 2019; Rojas-Rueda et al., 2019; Turner et al., 2016; Wellenius et al., 2017). Our simulation also relied on several strong assumptions about population mobility. Namely, we maintained state population constant and did not model other important movement considerations, such as community relationships (Gillespie, 2017). Finally, these results were premised on a large scale of population movement. While this may limit the feasibility of our projections, our goal was to examine the potential impacts of leveraging this policy intervention to its fullest extent, as one important piece of evidence for policymakers and as a blueprint for additional analyses. Our results are fully reproducible with available code to facilitate these extensions.

### 4.1. Conclusions

Our novel simulation-based health impact assessment demonstrates the importance of considering place-based policy approaches to desegregation as one mechanism to advance environmental health equity (Steil & Lens, 2023). Desegregation through equitable affordable housing development represents a structural remedy to structural injustice. This must be implemented in conjunction with people-based mechanisms and policies that improve environmental quality, particularly in communities impacted by environmental racism (Steil & Lens, 2023). These interventions lie at the nexus of social and environmental equity - further studies assessing the impacts of desegregation policy have the potential to catalyze lasting change.

## Data Availability

Data and programmatic code for analyses are available at https://github.com/CHEE-collaborative/CT-HIA and https://wonder.cdc.gov/.

https://github.com/CHEE-collaborative/CT-HIA

## Acknowledgements

Funding: This work was supported by the National Aeronautics and Space Administration [grant number 80NSSC22K1666] and the National Institutes of Health [grant numbers R01HL169171, R25ES029052]. The funding sources were not involved in study design, data analysis, interpretation of data, writing, or the decision to submit the article for publication.

