## Supplemental Materials for "Simulating desegregation through affordable housing development: an environmental health impact assessment of Connecticut zoning law"

**Figure A.1. Average exposure levels by Connecticut township in 2019.**

1.a. Population-weighted mean annual $PM_{2.5}$ (daily 24-hour mean) by township.

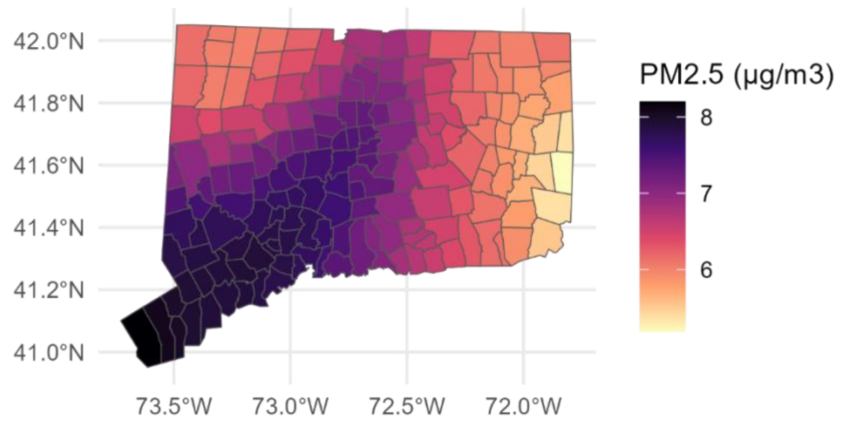


1.b. Population-weighted mean annual $O_3$ (daily 8-hour maximum) by township.

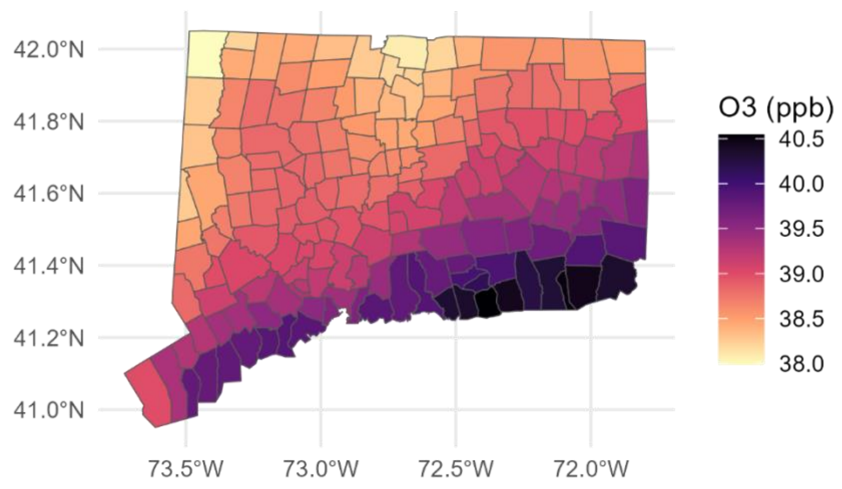


1.c. Population-weighted annual average surface-level $NO_2$ by township.

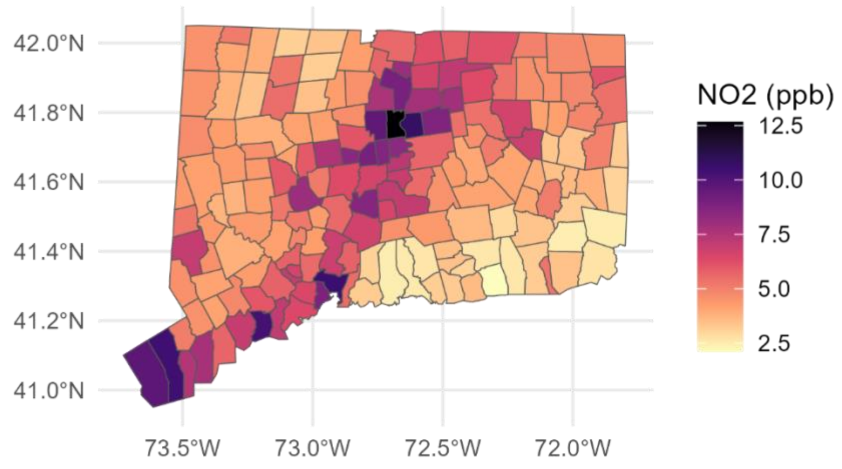


1.d. Population-weighted mean heat season (May 5–September 30) daily maximum heat index by township.

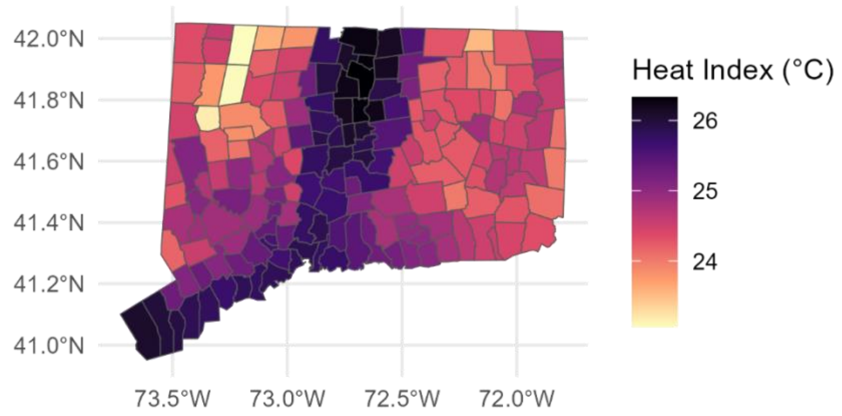


1.e. Mean annual normalized difference vegetation index (NDVI) by township.

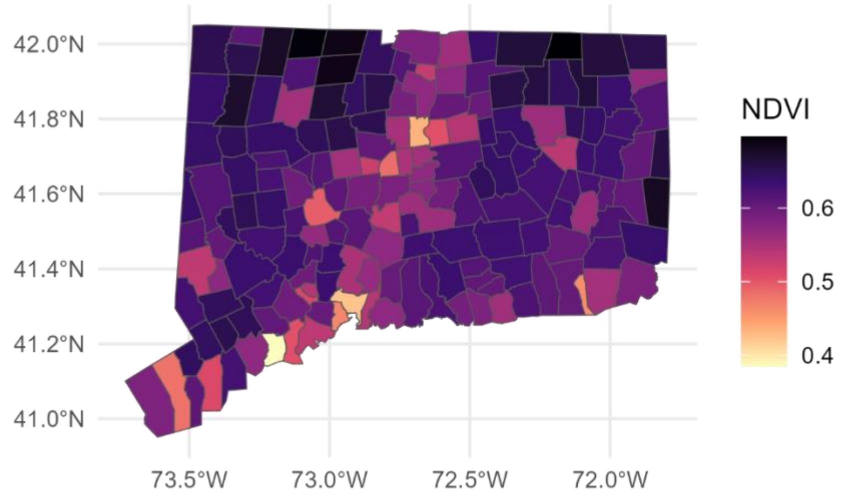


1.f. Mean annual road traffic noise by township.

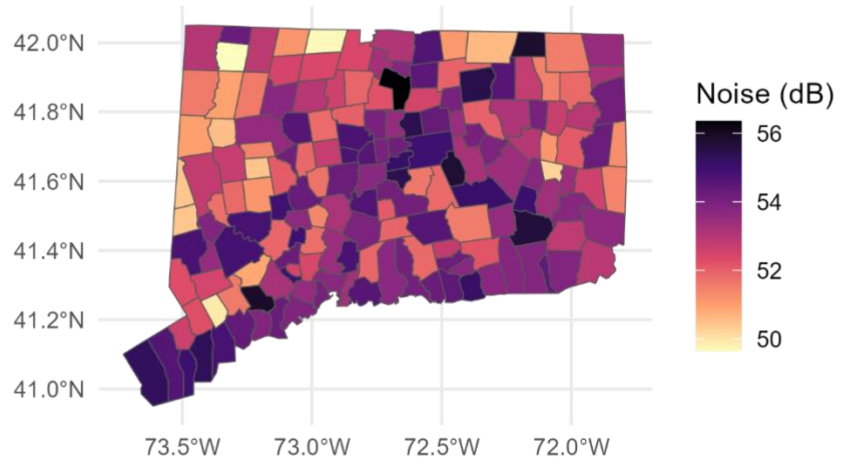

**Figure A.2. Affordable housing by Connecticut township in 2019.**

2.a. Percent of total housing units designated as affordable housing by township in 2019.

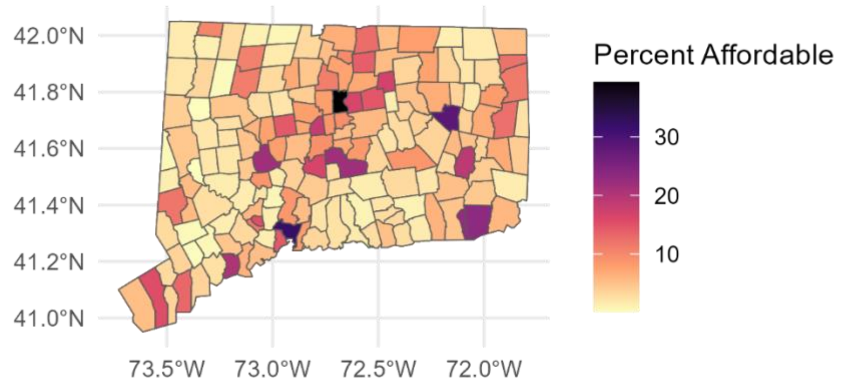


2.b. Townships with ≥10% housing units designated as affordable housing in 2019.

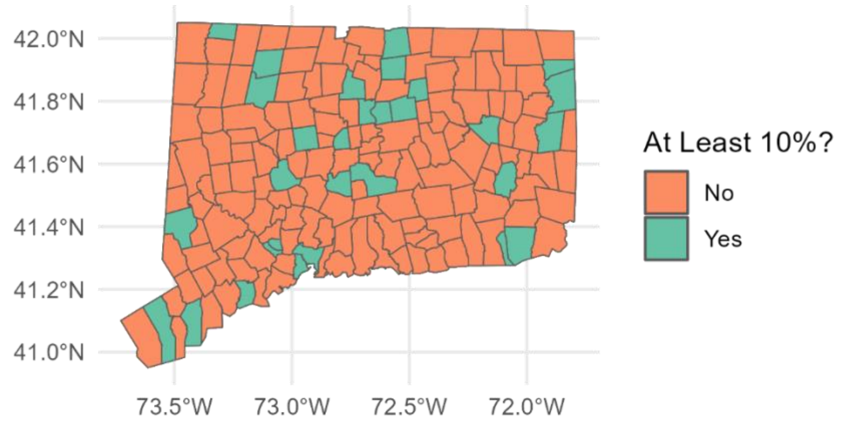

**Text A.1. Detailed methods for inverse distance weighting penalty**

The probability of moving between townships or staying within a township was weighted by the number of new housing units developed in the post-move township ($units_j$) divided by the total number of units developed across all townships ($\sum_{k=1}^{169} \text{units}_k$):

$$units_{prop} = \frac{units_j}{\sum_{k=1}^{169} units_k} \quad (1)$$

This probability of movement was also weighted by an inverse distance penalty. First, we computed the distance between population-weighted centroids of the pre-move and post-move township polygons ($distance_{j-i}$); in the case of same-township pairs, we computed half the distance between the two farthest points on the township boundary to approximate the township radius. We then calculated the inverse of this distance squared. The final inverse distance penalty was calculated as:

$$penalty_{j-i} = (\frac{1}{distance^2{}_{j-i}}) \quad (2)$$

We then multiplied the $units_{prop}$ term (equation 1) and $penalty_{j-i}$ term (equation 2) together. Finally, we divided this product by its sum across all township pairs before multiplying by the number of households in each pre-move town ($households_i$) to derive our final expected count (λ) of households moved between a given pair of townships. This final lambda can be shown as:

$$\lambda = \frac{\frac{units_{prop}}{penalty_{j-i}}}{\sum_{k=1}^{169} \frac{units_{prop}}{penalty_{k-i}}} * households_i \quad (3)$$

Our expected probability of movement for any household in a given pre/post-move township pair ($\lambda / households_i$) therefore summed to 1 across all pre-move townships, which assumed that each household either stayed in their pre-move township or moved to one of the other 168 townships in Connecticut.

**Text A.2. Detailed computation methods**

Census data were retrieved using the *tidycensus* package, daily temperature and vapor pressure were accessed using *daymetr*, Connecticut geospatial data were from *tigris*, and HUD income limits and town FIPS codes were from the *hudr* package (Hufkens et al., 2018; Richardson, 2022; Walker et al., 2024; Walker & Rudis, 2024). Daily maximum heat index was calculated using the *weathermetrics* package, and multi-group index of dissimilarity by county was calculated using the *OasisR* package (Anderson et al., 2013; Tivadar, 2019).

**Figure A.3. Map of low-income households across Connecticut townships at 2019 baseline versus simulated population distribution: Asian households.**

3.a. Asian low-income households at baseline in 2019.

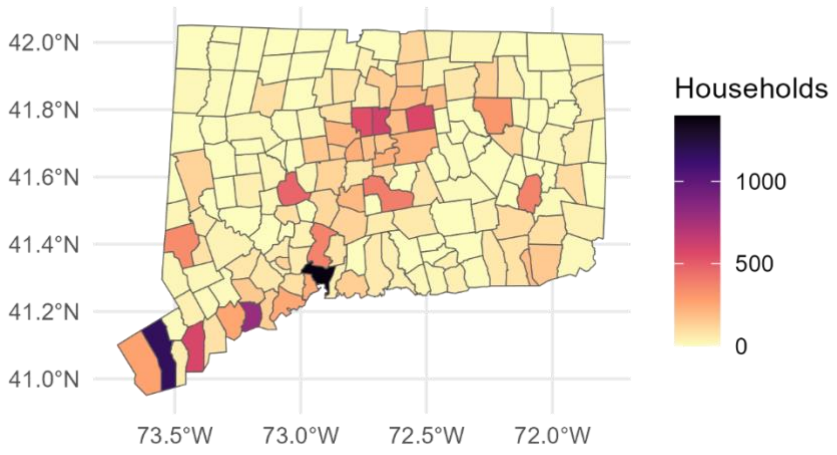


3.b. Asian low-income households after simulated desegregation.

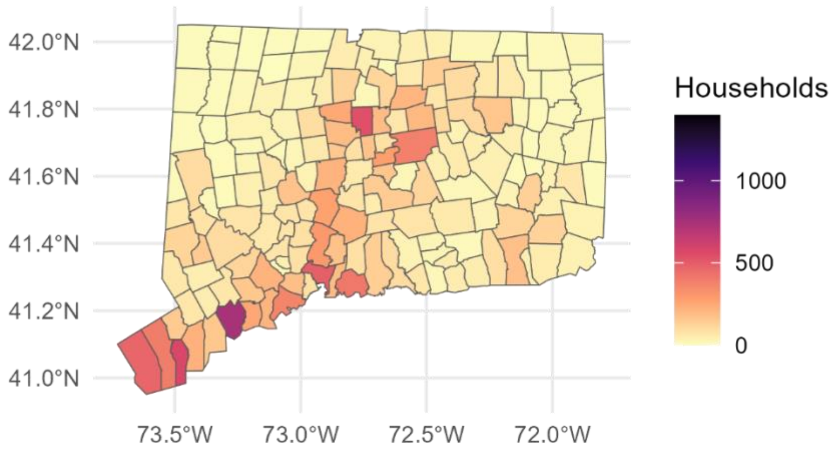

**Figure A.4. Map of low-income households across Connecticut townships at 2019 baseline versus simulated population distribution: non-Hispanic Black households.**

4.a. Non-Hispanic Black low-income households at baseline in 2019.

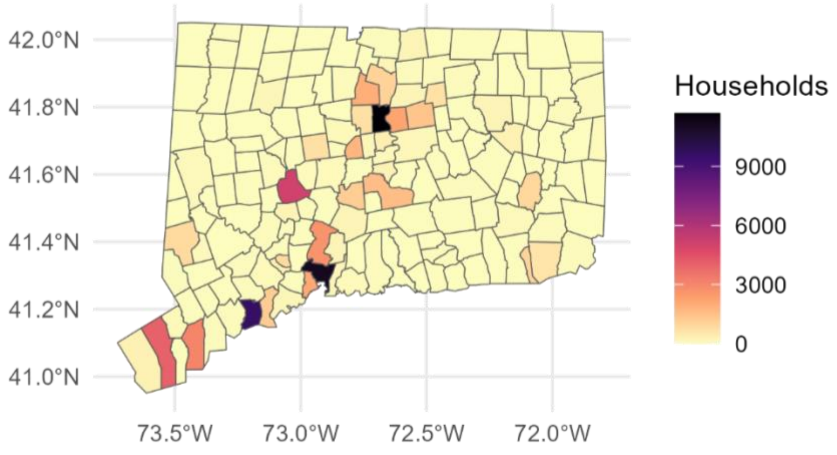


4.b. Non-Hispanic Black low-income households after simulated desegregation.

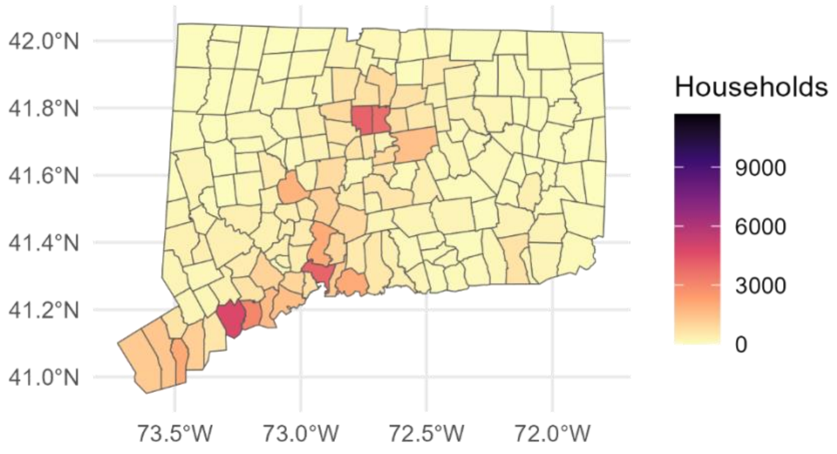

**Figure A.5. Map of low-income households across Connecticut townships at 2019 baseline versus simulated population distribution: Hispanic or Latino households.**

5.a. Hispanic or Latino low-income households at baseline in 2019.

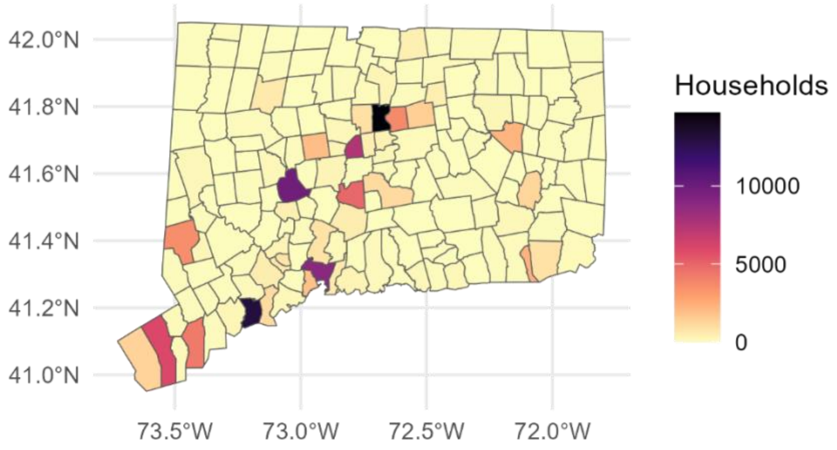


5.b. Hispanic or Latino low-income households after simulated desegregation.

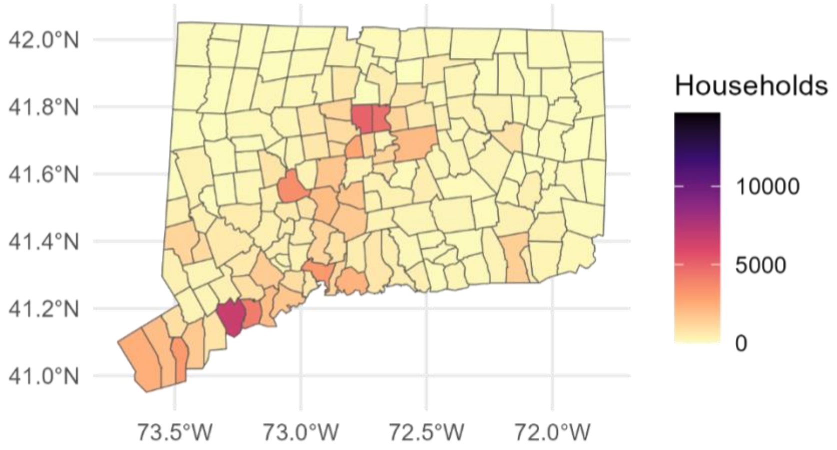

**Figure A.6. Map of low-income households across Connecticut townships at 2019 baseline versus simulated population distribution: non-Hispanic White households.**

6.a. Non-Hispanic White low-income households at baseline in 2019.

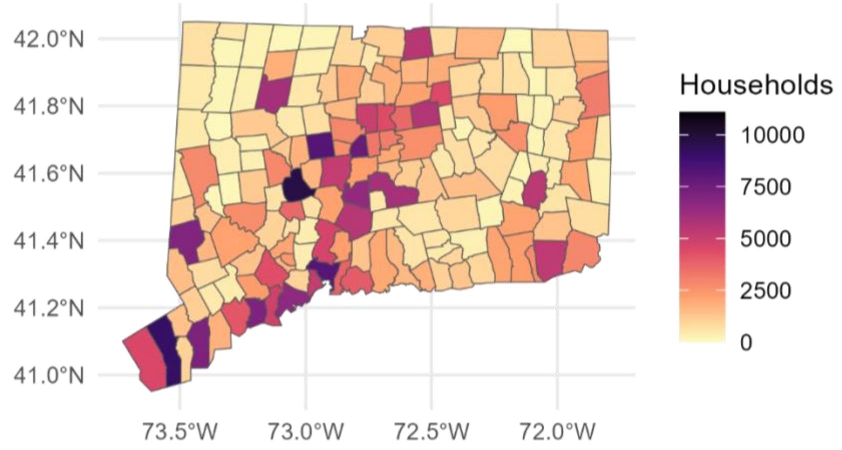


6.b. Non-Hispanic White low-income households after simulated desegregation.

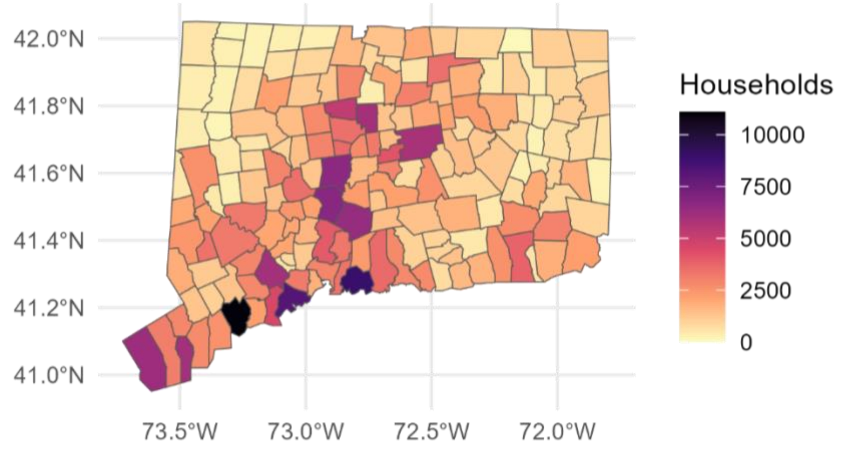

**Table A.1. Multi-group index of dissimilarity by county, at baseline and post-simulation average in 2019.**

| County | Baseline | Post-simulation |
|---|---|---|
| Fairfield | 0.39 | 0.35 |
| Hartford | 0.45 | 0.4 |
| Litchfield | 0.27 | 0.2 |
| Middlesex | 0.36 | 0.26 |
| New Haven | 0.41 | 0.37 |
| New London | 0.36 | 0.31 |
| Tolland | 0.31 | 0.24 |
| Windham | 0.5 | 0.46 |

**Figure A.7. Map of multi-group index of dissimilarity by county, (A) at baseline and (B) post-simulation average in 2019.**

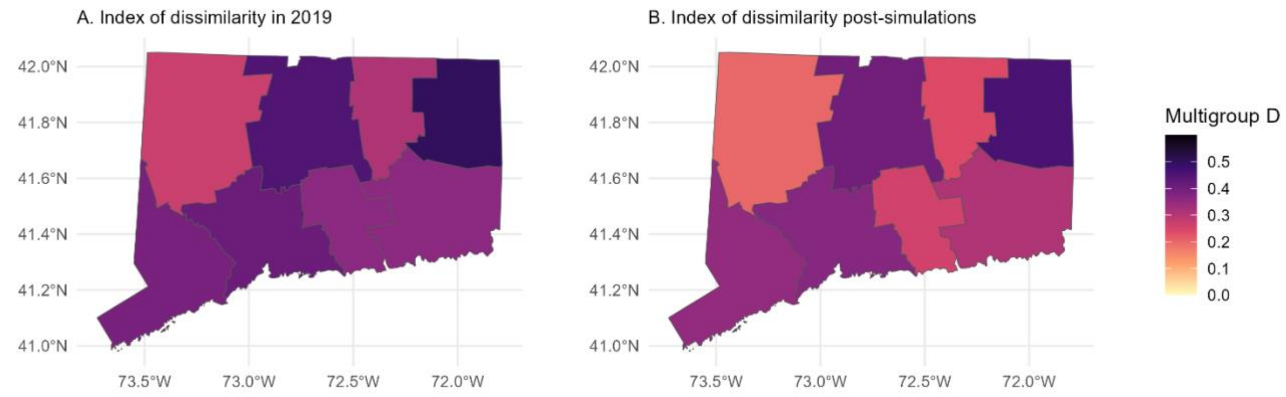

**Table A.2. Average exposure levels among low-income households at baseline and post-simulation based on average town-level exposures in 2019** (mean town-level exposure level with first and third quartiles in baseline and average post-simulation population distribution).

| Exposure | Pre-simulation mean (Q1, Q3) | Post-simulation mean (Q1, Q3) |
|---|---|---|
| $PM_{2.5}$ (µg/m$^3$) | 7.28 (7, 7.73) | 7.29 (6.98, 7.79) |
| $O_3$ (ppb) | 39.13 (38.68, 39.47) | 39.17 (38.76, 39.72) |
| $NO_2$ (ppb) | 7.46 (5.49, 9.65) | 6.21 (4.36, 7.48) |
| Heat index (°C) | 25.46 (25.07, 25.86) | 25.37 (24.87, 25.82) |
| Greenness (NDVI) | 0.53 (0.48, 0.60) | 0.58 (0.55, 0.62) |
| Noise (dB) | 53.88 (53.53, 54.58) | 53.70 (53.1, 54.58) |

NDVI, normalized difference vegetation index

**Table A.3. Annual deaths averted after simulated desegregation by ethnoracial group, without inverse distance weighting**

| Ethnoracial Group | Heat | NDVI | $NO_2$ | Noise | $O_3$ | $PM_{2.5}$ |
|---|---|---|---|---|---|---|
| Asian | 0 (0, 0) | 2 (1, 2) | 1 (1, 1) | 0 (0, 0) | 0 (0, 0) | 0 (0, 0) |
| Hispanic/Latino | 0 (0, 1) | 27 (13, 40) | 12 (8, 17) | 2 (1, 3) | 0 (0, 0) | 2 (1, 2) |
| Non-Hispanic Black | 1 (0, 1) | 43 (21, 66) | 21 (14, 28) | 2 (1, 4) | 0 (0, 0) | 3 (2, 4) |
| Non-Hispanic White | 1 (0, 2) | 107 (51, 163) | 50 (33, 67) | 10 (4, 16) | -1 (-2, -1) | -2 (-3, -1) |
| Total | 3 (1, 4) | 179 (85, 272) | 84 (56, 112) | 14 (6, 22) | -2 (-2, -1) | 2 (2, 3) |

NDVI, normalized difference vegetation index

**Table A.4. Annual deaths averted after simulated desegregation by ethnoracial group, 2018**

| Ethnoracial Group | Heat | NDVI | $NO_2$ | Noise | $O_3$ | $PM_{2.5}$ |
|---|---|---|---|---|---|---|
| Asian | 0 (0, 0) | 2 (1, 2) | 1 (0, 1) | 0 (0, 0) | 0 (0, 0) | 0 (0, 0) |
| Hispanic/Latino | 0 (0, 0) | 25 (13, 38) | 10 (7, 13) | 1 (0, 2) | 0 (0, 0) | 0 (0, 1) |
| Non-Hispanic Black | 0 (0, 1) | 40 (20, 60) | 17 (11, 22) | 1 (0, 2) | 0 (0, 0) | 1 (0, 1) |
| Non-Hispanic White | 1 (0, 2) | 115 (58, 172) | 44 (30, 58) | 9 (4, 14) | -1 (-1, 0) | -2 (-3, -2) |
| Total | 1 (0, 2) | 182 (92, 272) | 71 (49, 94) | 11 (4, 17) | -1 (-1, -1) | -1 (-2, -1) |

NDVI, normalized difference vegetation index
